# Discordance Between Perceived Health Information Competence and Cancer Prevention Knowledge in U.S. Adults: A Cross-Sectional Study

**DOI:** 10.64898/2026.05.28.26354370

**Authors:** Cooper Lee, Andrew Wong, Luke Yin, Yoo Choi

**Author notes:** **Corresponding author:** Cooper Lee.

## Abstract

**Background:** Self-reported confidence in health information seeking does not reliably predict accurate health knowledge, yet the population-level distribution of this discordance and its demographic predictors have received limited direct study. This study aimed to identify and characterize a Confident-Incorrect phenotype among U.S. adults: individuals with high perceived health information competence who simultaneously hold inaccurate or fatalistic beliefs about cancer.

**Methods:** Cross-sectional analysis of HINTS 7 (*N* = 7,278). A Confidence Index (3-item digital literacy composite; Cronbach’s *α* = 0.674) and an Evidence-Consistent Knowledge Score (factual cancer knowledge minus a cancer fatalism composite; fatalism subscale *α* = 0.563) were computed and combined into a discordance framework. Median-split classification produced four phenotypes. Gaussian Mixture Model clustering with four components provided moderate independent validation (inter-method agreement = 65.2%). Survey-weighted multinomial logistic regression (*n* = 5,771; McFadden pseudo-*R*^2^ = 0.129) examined phenotype predictors.

**Results:** An estimated 20.3% of U.S. adults were classified as Confident-Incorrect. They reported confidence levels similar to Well-Informed adults (*z* = 0.72 vs. 0.82) but scored 2.8-fold lower on objective cancer knowledge (0.74 vs. 2.06 out of 4) and exhibited the highest cancer fatalism of any phenotype (3.17 vs. 1.65 out of 4). Only 14.3% correctly identified alcohol as a cancer risk factor (vs. 58.8% of Well-Informed adults). Cancer screening rates did not differ meaningfully across phenotypes. Lower education (OR = 0.754), Hispanic ethnicity (OR = 1.788), non-Hispanic Black race (OR = 1.893), higher social media use (OR = 1.097), and lower trust in scientists (OR = 0.749) independently predicted Confident-Incorrect membership.

**Conclusions:** An estimated one in five U.S. adults is overconfident in health information competence while holding substantially inaccurate beliefs about cancer prevention. Cancer screening rates did not follow the expected gradient across phenotypes, a null finding that cautions against inferring immediate behavioral impact from observed belief gaps. Interventions targeting specific factual errors and cancer fatalism are more likely to reach this group than general health literacy programs.

## 1 Background

Most U.S. adults seek health information online, and a growing share encounter it primarily through social media [1–3]. The problem is not limited access to information; it is that confidence in finding information does not indicate that the information found is accurate. eHealth literacy research has documented that perceived information competence and objective accuracy often diverge [4, 5]: the Dunning–Kruger effect describes how people with limited domain knowledge overestimate their own competence due to insensitivity to their deficits [6], and evidence on belief correction indicates that people who are confident in their existing beliefs are also the most resistant to subsequent correction [7, 8]. Individuals who recognize a knowledge gap may be more inclined to seek professional guidance; those who perceive themselves as already well-informed face less incentive to do so [6].

Cancer prevention makes this problem concrete. Cancer risk knowledge among U.S. adults is limited [9], including for well-documented factors such as alcohol [10] and hepatitis B and C viruses [11], and many Americans hold fatalistic beliefs about cancer that are directly incompatible with prevention behavior [12, 13]. Health misinfor-mation circulating on social media has been associated with lower cancer screening rates [14] and eroded trust in public health institutions [15]. Yet no nationally representative study has directly examined whether people who perceive themselves as most capable of navigating health information are in fact better informed about cancer risk, or whether some are confidently misinformed.

This study uses the Health Information National Trends Survey, Cycle 7 (HINTS 7, 2024) to address that gap. We compute a discordance score separating perceived health information competence from evidence-consistent cancer knowledge, classify U.S. adults into four phenotypes based on that discordance, and identify the demographic and behavioral predictors of Confident-Incorrect status.

## 2 Methods

### 2.1 Dataset

HINTS 7 is a nationally representative cross-sectional survey of U.S. adults administered by the National Cancer Institute (NCI) from February through June 2024 (*N* = 7,278) [16, 17], using probability-based address sampling with mail distribution in English and Spanish. Survey weights (PERSON FINWT0 through PER-SON FINWT50) and complex design variables (STRA-TUM, VAR CLUSTER) were applied throughout. The dataset is publicly available and de-identified; this analysis is exempt from institutional review board (IRB) review under 45 CFR 46.101(b)(4) [16].

### 2.2 Measures

#### Confidence Index

Three HINTS 7 items assessed digital health literacy: ability to find health information online without help, self-rated quality of health information search skills, and how frustrating health information searching is. Items were reverse-coded where needed so that higher values indicate greater confidence, then averaged and standardized (*M* = 0, *SD* = 1; Cronbach’s *α* = 0.674).

##### Evidence-Consistent Knowledge Score

A Knowledge Score (0 to 4) summed binary correct/incorrect responses for four factual items: whether ‘1 in 100’ is greater than ‘1 in 1,000’ (cancer numeracy) [18]; whether alcohol increases cancer risk [10]; whether hepatitis B virus (HBV) can cause cancer; and whether hepatitis C virus (HCV) can cause cancer [11]. A Cancer Fatalism Score (0 to 4) summed binary endorsement of four items from validated HINTS fatalism scales [12, 13]: ‘everything causes cancer’; ‘not much you can do to lower your chances of getting cancer’; ‘cancer is so serious you might as well not try to prevent it’; and ‘too many recommendations to know which to follow.’ Internal consistency for the fatalism subscale was below the conventional threshold (Cronbach’s *α* = 0.563), reflecting genuine heterogeneity across fatalistic belief domains; this composite should be interpreted with appropriate caution. Both scores were standardized and combined (*z*_knowledge_*− z*_fatalism_), with higher values indicating greater factual accuracy and less fatalism.

##### Discordance Score and Phenotype Classification

A Discordance Score was computed as *z*_confidence_*−z*_evidence_. At the population level, the two components were weakly but positively correlated (*r* = 0.227, *p <* .001), indicating that more confident adults tended to be somewhat more knowledgeable on average. The overconfidence pattern of interest exists in the tail of that distribution, not as a population-level inverse. Participants with complete data on both scores (*n* = 6,608; 90.8%) were assigned to four phenotypes by median split: Well-Informed (above-median on both), Confident-Incorrect (above-median confidence, below-median knowledge), Uncertain-Learner (below-median confidence, above-median knowledge), and Disengaged (below-median on both). Gaussian Mixture Model (GMM) clustering with four components and 20 random initializations provided moderate independent empirical validation of the group structure [19].

### 2.3 Statistical Analysis

Weighted prevalence estimates and group means were computed using final person-level weights. Survey-weighted multinomial logistic regression with Well-Informed as the reference category was conducted among the *n* = 5,771 classified respondents with complete data on all covariates (837 of the 6,608 classified individuals were excluded due to missing values on income [*n*_missing_ = 820] and/or race/ethnicity [*n*_missing_ = 760], with overlap). Covariates included age (centered at 50 years), sex, education (ordinal 1 to 7), race/ethnicity (vs. non-Hispanic White), household income (ordinal), internet use frequency, social media visit frequency, physician trust, and scientist trust. All analyses used Python 3.10 (statsmodels v0.14, scikit-learn v1.3).

### 2.4 Use of Artificial Intelligence Tools

AI language tools were used to assist with manuscript drafting and editing. The authors take full responsibility for the accuracy of all data, analyses, and interpretations presented.

## 3 Results

### 3.1 Phenotype prevalence

Of 7,278 respondents, 6,608 (90.8%) were classified. Weighted national estimates: Well-Informed 37.1%, Confident-Incorrect 20.3%, Uncertain-Learner 19.6%, Disengaged 23.0% (Fig. 1). GMM validation yielded cluster centroids consistent with the four theoretical quadrants and moderate inter-method agreement with the median-split labels (65.2%). The moderate agreement confirms a four-group structure in the data while reflecting the inherent arbitrariness of dichotomous cutoffs applied to continuous variables.

**Figure 1:**
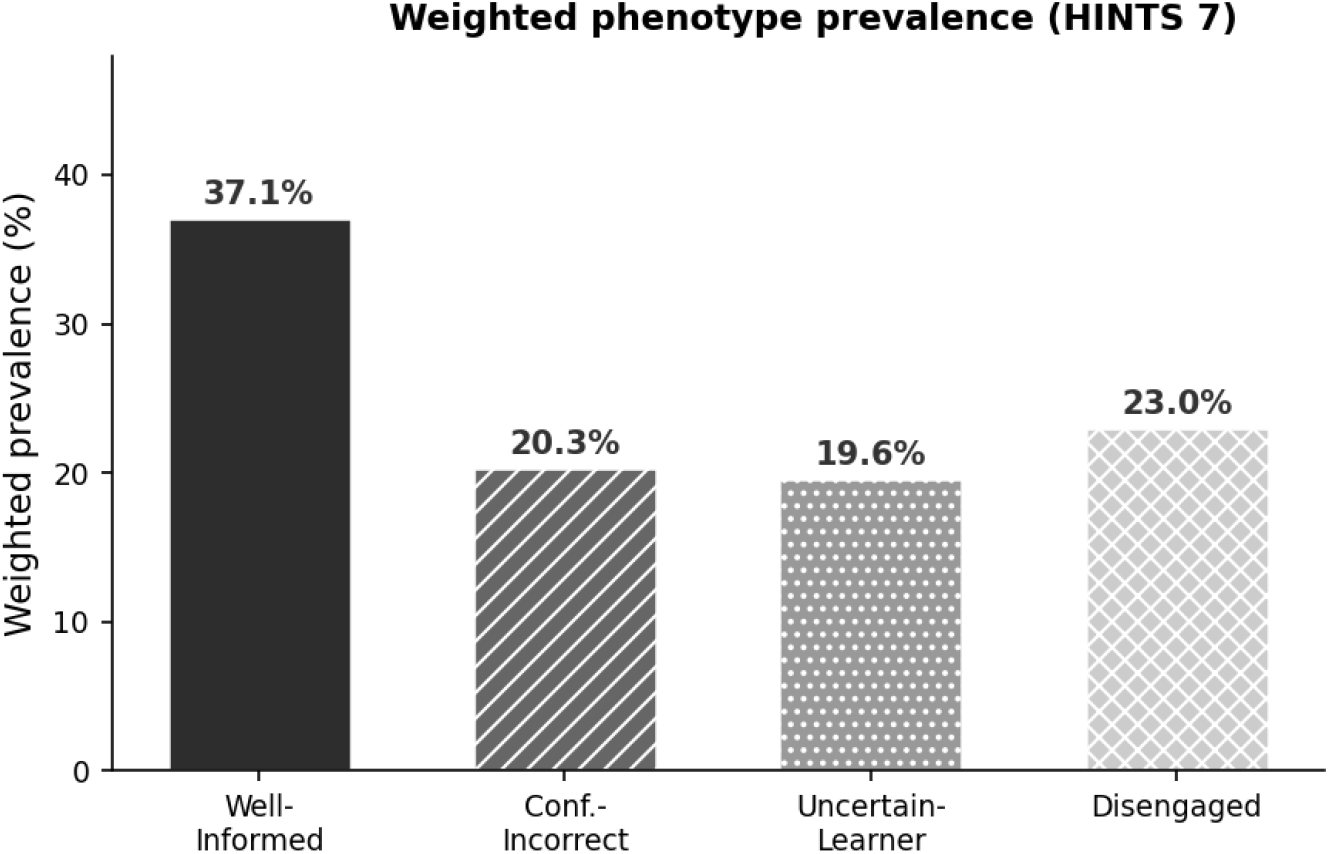
Weighted phenotype prevalence among U.S. adults (HINTS 7, *N* = 7,278). An estimated 20.3% of U.S. adults are Confident-Incorrect.

### 3.2 Knowledge, beliefs, and group profiles

Table 1 presents weighted characteristics for each phenotype. The Confident-Incorrect group had a confidence *z*-score comparable to the Well-Informed group (0.72 vs. 0.82) but scored 2.8-fold lower on objective cancer knowledge (0.74 vs. 2.06 out of 4) and recorded the highest cancer fatalism of all four groups (3.17 vs. 1.65 out of 4; all comparisons *p <* .001). Fig. 2 illustrates this separation: the two groups overlap substantially on the confidence axis but diverge sharply on the knowledge axis.

**Table 1:** Weighted descriptive characteristics by health information phenotype (HINTS 7, *n* = 6,608)

|  | Well-Informed | Confident-Incorrect | Uncertain-Learner | Disengaged |
| --- | --- | --- | --- | --- |
| $n$ (unweighted) | 2,333 | 1,191 | 1,486 | 1,598 |
| Weighted prevalence (%) | 37.1 | 20.3 | 19.6 | 23.0 |
| Age (mean, years) | 44.7 | 43.7 | 54.5 | 54.7 |
| Knowledge Score, mean (0–4) | 2.06 | 0.74 | 1.89 | 0.61 |
| Fatalism Score, mean (0–4) | 1.65 | 3.17 | 1.75 | 3.14 |
| Confidence Index, mean $z$-score | 0.82 | 0.72 | −0.78 | −0.85 |
| Trust in scientists, mean (1–4) | 3.59 | 3.36 | 3.32 | 3.04 |
| Social media visits, mean (1–5) | 4.42 | 4.55 | 3.64 | 3.75 |
| ‘Everything causes cancer’ (%) | 57.7 | 87.7 | 53.6 | 85.2 |
| Cancer numeracy correct (%) | 89.7 | 57.0 | 81.0 | 43.2 |
| Alcohol–cancer link correct (%) | 58.8 | 14.3 | 51.7 | 11.1 |
| Knows HBV causes cancer (%) | 30.0 | 1.8 | 30.4 | 2.9 |
| Knows HCV causes cancer (%) | 28.4 | 1.2 | 26.4 | 3.7 |
| Prevention possible (%) | 91.8 | 46.4 | 84.5 | 41.1 |
| Colorectal screening (%)<sup>a</sup> | 41.2 | 37.5 | 47.7 | 44.6 |
| Current smoker (%) | 8.3 | 11.0 | 14.1 | 17.4 |

All estimates survey-weighted. Between-group differences on continuous variables: $F$ -test $p < .001$ .
<sup>a</sup> Screening rates did not follow the expected gradient across phenotypes (see text).

**Figure 2:**
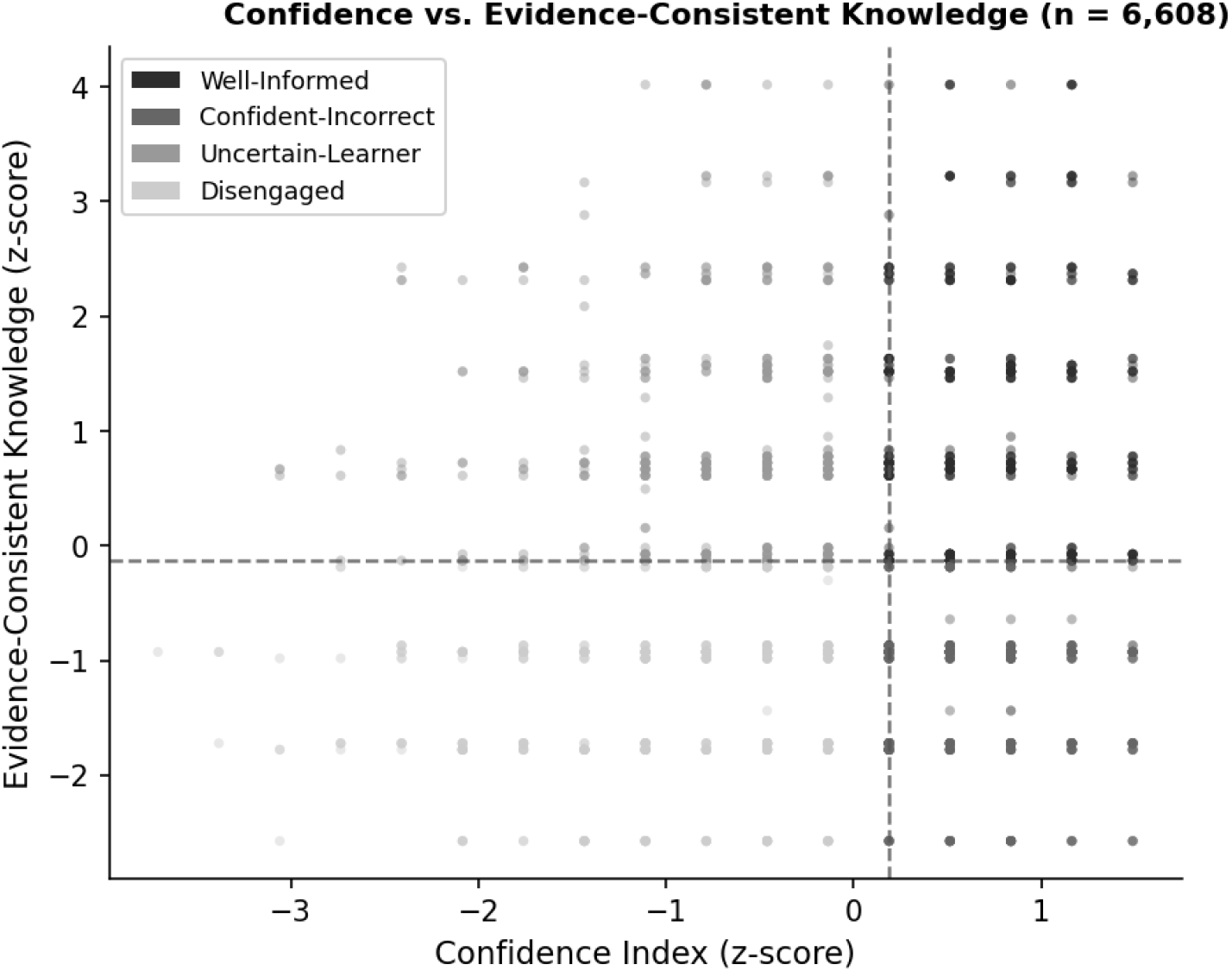
Confidence Index vs. Evidence-Consistent Knowledge Score (*n* = 6,608). Dashed lines indicate median thresholds. Confident-Incorrect (medium grey) and Well-Informed (dark) overlap substantially on confidence but diverge sharply on knowledge.

Item-level results are presented in Fig. 3. Only 14.3% of Confident-Incorrect adults correctly identified alcohol as a cancer risk factor, compared with 58.8% of Well-Informed adults. The numeracy item was answered correctly by 57.0% of Confident-Incorrect adults vs. 89.7% of Well-Informed adults. Fewer than 2% of Confident-Incorrect adults knew that HBV or HCV can cause cancer, compared with 28 to 30% of Well-Informed adults. On fatalism items, 87.7% of Confident-Incorrect adults endorsed ‘everything causes cancer’ and 53.6% did not believe cancer prevention is possible; both figures closely parallel those of the Disengaged group (85.2% and 58.9%), despite Confident-Incorrect adults reporting confidence levels as high as the Well-Informed group, among whom only 8.2% endorsed the belief that prevention is not possible. Social media use was highest in the Confident-Incorrect group (mean 4.55 out of 5 vs. 4.42 for Well-Informed), and scientist trust was lower (3.36 vs. 3.59 out of 4).

**Figure 3:**
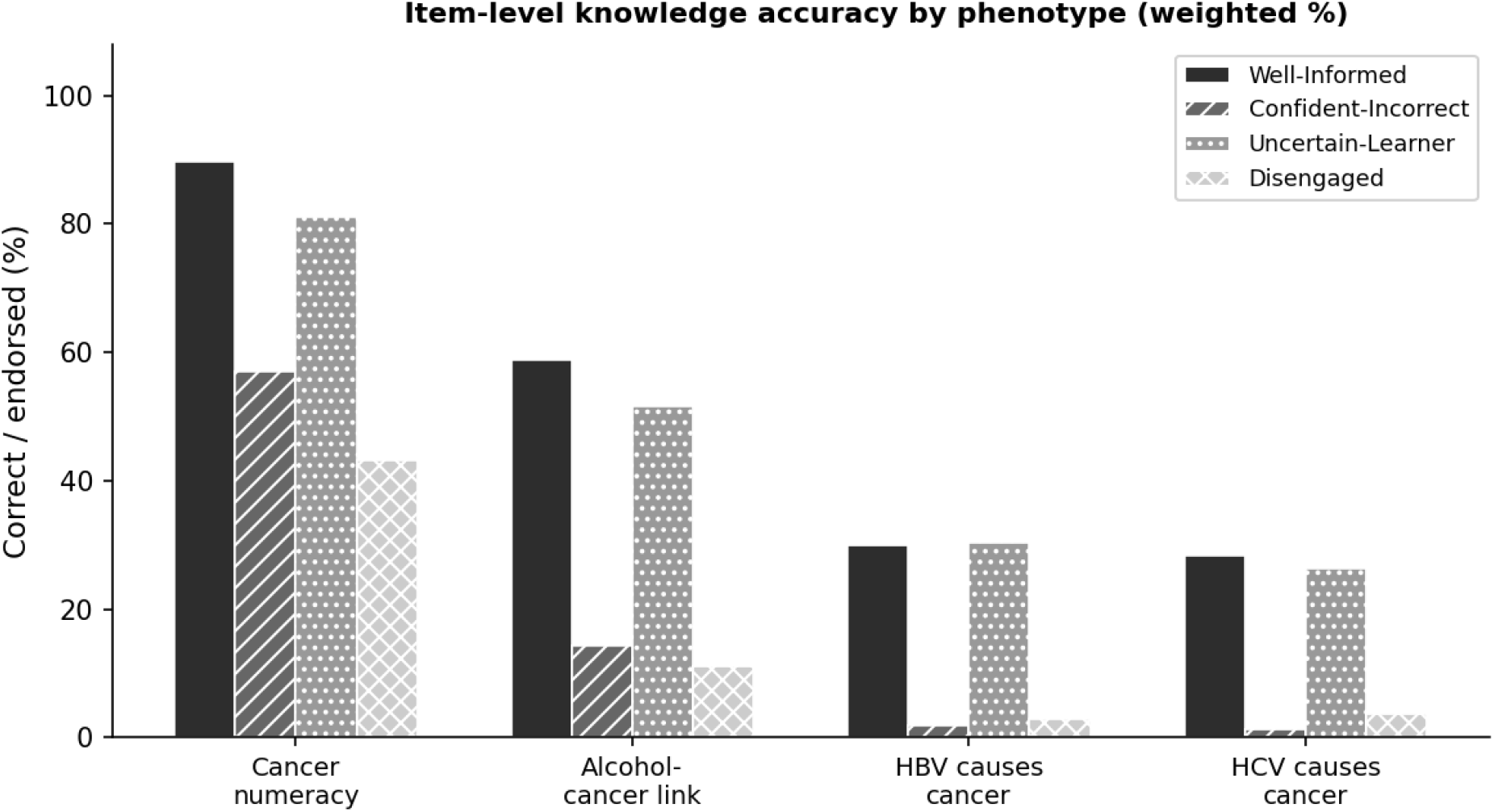
Item-level knowledge and belief accuracy by phenotype (weighted %). Confident-Incorrect (medium grey) consistently underperforms Well-Informed (dark) across all items despite equivalent confidence.

### 3.3 Behavioral outcomes

Colorectal cancer screening rates were unexpectedly higher in the Uncertain-Learner (47.7%) and Disengaged (44.6%) groups than in the Well-Informed (41.2%) and Confident-Incorrect (37.5%) groups. Provider visit frequency was similar across all groups (means 2.79 to 3.01). The knowledge and belief differences observed across phenotypes did not translate into measurable behavioral differences in this cross-sectional sample.

### 3.4 Predictors of phenotype membership

Table 2 and Fig. 4 present regression results. Education was the strongest predictor across all non-reference phenotypes: each one-unit increase reduced the odds of Confident-Incorrect classification by 24.6% (OR = 0.754 [0.710–0.802], *p <* .001). Hispanic adults (OR = 1.788 [1.463–2.185], *p <* .001) and non-Hispanic Black adults (OR = 1.893 [1.525–2.348], *p <* .001) were significantly more likely to be Confident-Incorrect than non-Hispanic White adults after adjusting for education and income. Social media visit frequency positively predicted Confident-Incorrect membership (OR = 1.097 [1.023–1.177], *p* = .009); general internet use frequency did not (*p* = .077). Lower scientist trust predicted Confident-Incorrect status (OR = 0.749 [0.672–0.834], *p <* .001). Physician trust (*p* = .400), age (*p* = .695), sex (*p* = .396), and income (*p* = .083) were not significant.

**Table 2:** Survey-weighted multinomial logistic regression: predictors of phenotype membership (reference = Well-Informed; *n* = 5,771; pseudo-*R*^2^ = 0.129)

| Predictor | Confident-Incorrect |  | Uncertain-Learner |  | Disengaged |  |
| --- | --- | --- | --- | --- | --- | --- |
|  | OR | 95% CI | OR | 95% CI | OR | 95% CI |
| Age (50 yrs ctr.) | 1.00 | [0.994, 1.004] | 1.04*** | [1.030, 1.040] | 1.04*** | [1.029, 1.040] |
| Female sex | 0.93 | [0.796, 1.094] | 1.10 | [0.938, 1.281] | 1.13 | [0.963, 1.333] |
| Education level | 0.754*** | [0.710, 0.802] | 0.778*** | [0.732, 0.826] | 0.647*** | [0.609, 0.688] |
| Hispanic | 1.788*** | [1.463, 2.185] | 1.130 | [0.910, 1.404] | 1.333* | [1.070, 1.661] |
| Non-Hispanic Black | 1.893*** | [1.525, 2.348] | 0.682** | [0.535, 0.870] | 1.053 | [0.834, 1.328] |
| Non-Hispanic Asian | 0.960 | [0.663, 1.389] | 1.795*** | [1.309, 2.461] | 1.404 | [0.970, 2.033] |
| Household income | 0.965 | [0.926, 1.005] | 0.862*** | [0.828, 0.897] | 0.847*** | [0.813, 0.882] |
| Internet use freq. | 0.861 | [0.730, 1.016] | 0.591*** | [0.518, 0.675] | 0.552*** | [0.484, 0.630] |
| Social media visits | 1.097** | [1.023, 1.177] | 0.889*** | [0.839, 0.941] | 0.922** | [0.868, 0.980] |
| Trust in doctor | 1.064 | [0.921, 1.229] | 0.811** | [0.706, 0.932] | 0.851* | [0.741, 0.977] |
| Trust in scientists | 0.749*** | [0.672, 0.834] | 0.888* | [0.796, 0.990] | 0.663*** | [0.596, 0.737] |

Reference category = Well-Informed. OR = odds ratio. \* $p < .05$ , \*\* $p < .01$ , \*\*\* $p < .001$ . Non-significant for Confident-Incorrect: age, sex, Asian race, income, internet use, physician trust.

**Figure 4:**
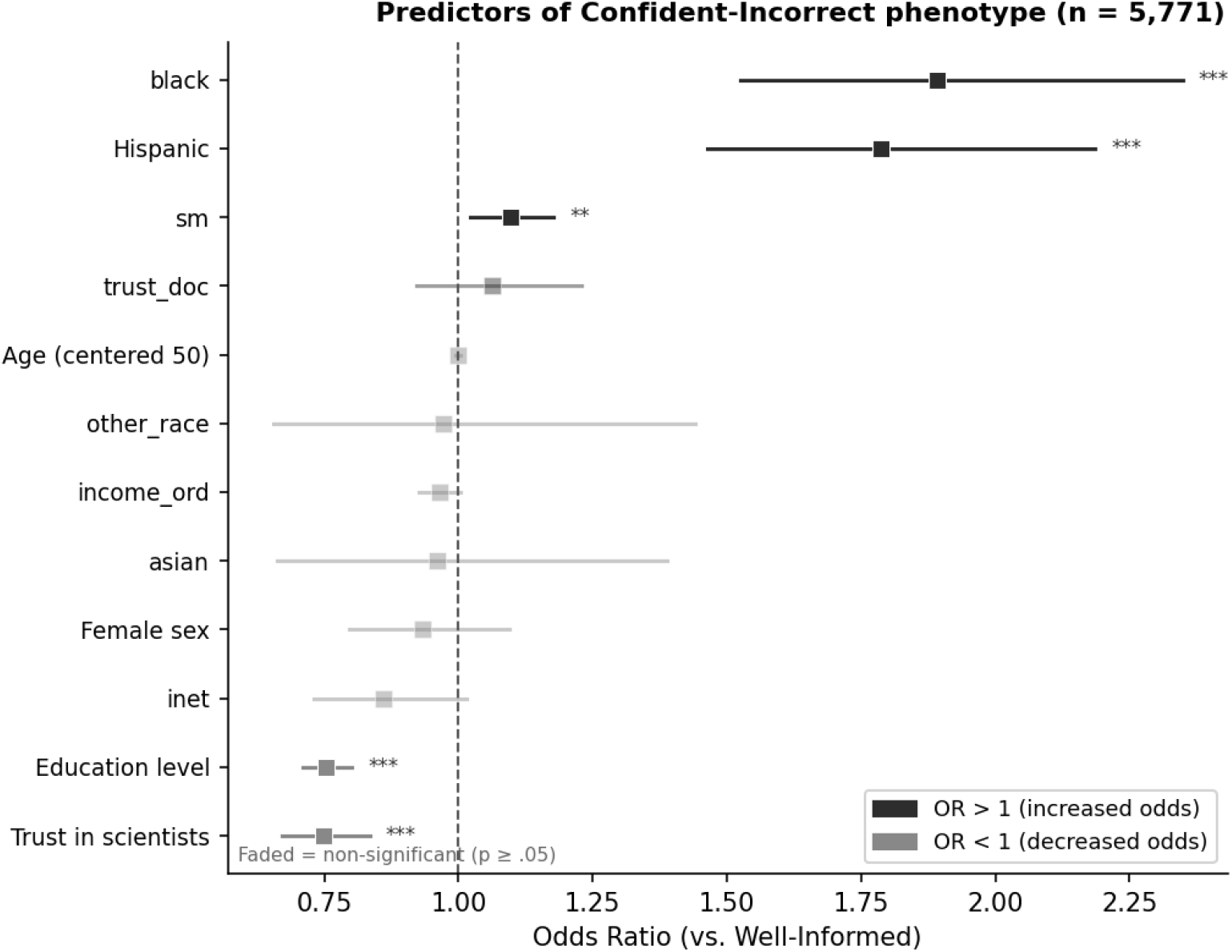
Forest plot: predictors of Confident-Incorrect vs. Well-Informed (*n* = 5,771). Dark fill = OR *>* 1; light fill = OR *<* 1. Faded markers = non-significant. ^*\**^*p <* .05, ^*\*\**^*p <* .01, ^*\*\*\**^*p <* .001.

## 4 Discussion

The Confident-Incorrect group is not simply a low-knowledge group. Its members report confidence levels comparable to the most accurate adults in the sample, which is the core problem: when individuals do not recognize a gap in their own knowledge, there is no internal prompt to seek correction [6, 7]. Cancer beliefs in this group were nearly indistinguishable from those of the Disengaged group, the least-engaged population segment, despite confidence levels comparable to the most accurate group. The finding that social media visit frequency, but not general internet use frequency, predicted Confident-Incorrect membership suggests that the specific information environment on social media platforms, where emotionally resonant content spreads regardless of accuracy [2, 15], is a more important driver than digital engagement in general.

Scientist trust was lower among Confident-Incorrect adults than Well-Informed adults, but physician trust did not differ significantly. That institutional credibility was affected while personal clinical trust was not is consistent with prior evidence on the differential effects of health misinformation on trust in online versus personal sources [5, 15]. Primary care encounters therefore remain a viable corrective channel for this group. The observed racial and ethnic disparities are consistent with structural communication inequalities documented elsewhere [20]. Hispanic and non-Hispanic Black adults were significantly more likely to be Confident-Incorrect after controlling for education and income; this residual disparity likely reflects differential exposure to health misinformation in culturally targeted media [2], higher rates of cancer fatalism documented in these communities [13, 21], histories of medical mistreatment that undermine institutional health messaging [22], and reduced access to evidence-based health education [9]. Effective interventions will likely require culturally tailored approaches developed in collaboration with these communities [20].

Cancer screening rates were higher in the Uncertain-Learner and Disengaged groups than in the Well-Informed and Confident-Incorrect groups, the reverse of the expected gradient. This pattern most plausibly reflects the predominant role of clinician recommendation and insurance coverage in determining screening uptake: structural factors that operate largely independently of information phenotype in a cross-sectional sample. Whether phenotype membership predicts screening behavior longitudinally is a question this study cannot address.

Because the Confident-Incorrect group already perceives itself as informed, general health literacy programs [23–25] are unlikely to be effective. The challenge is not teaching people to locate and evaluate health information but correcting the specific factual errors they already hold with confidence: that alcohol raises cancer risk [10], that hepatitis viruses can cause cancer [11], and that fatalistic beliefs about cancer prevention are inconsistent with current evidence [12, 13]. Prebunking approaches (which preemptively expose individuals to attenuated versions of common misleading arguments before full exposure [26– 28]) may be better suited to this group than post-hoc corrections. Given the intact physician trust observed among Confident-Incorrect adults, primary care encounters represent a viable and potentially high-yield channel for delivering targeted factual corrections.

### 4.1 Limitations

The cross-sectional design precludes causal inference. The Knowledge and Fatalism scores are constructed from available HINTS items rather than validated psychometric batteries; the fatalism subscale’s internal consistency (Cronbach’s *α* = 0.563) was below the conventional threshold and the composite should be interpreted accordingly. The 65.2% GMM–median-split agreement is moderate rather than strong, indicating a four-group structure in the data but also that exact boundaries are somewhat arbitrary. Missing data for income (12.4%) and race/ethnicity (11.5%) may introduce selection bias. HINTS 7 does not identify which social media platforms respondents use most frequently.

## 5 Conclusions

An estimated one in five U.S. adults combines high perceived health information competence with substantially incorrect beliefs about cancer prevention. This group actively seeks health information, primarily through social media, and is confident in its own assessment of that information. Cancer screening rates did not follow the expected gradient across phenotypes in this cross-sectional sample, a null finding that cautions against inferring immediate behavioral impact from observed belief gaps. The knowledge deficits, however, are large and consistent across multiple items. Perceived competence in health information seeking does not reliably indicate accuracy, and interventions premised on high self-reported eHealth literacy as a proxy for well-informed beliefs are unlikely to reach the individuals most in need of correction.

## Data Availability

All data used in this study are publicly available from the National Cancer Institute's Health Information National Trends Survey (HINTS) repository at https://hints.cancer.gov/data/download-data.aspx. No new data were generated. All analytical code and derived outputs are available upon reasonable request to the corresponding author.

## Author Contributions

C.L. conceptualized the study, performed all data curation, formal analysis, and methodology, and drafted the manuscript. A.W. contributed to the methodology and reviewed and edited the manuscript. L.Y. reviewed and edited the manuscript. Y.C. provided critical revisions and reviewed and edited the manuscript. All authors read and approved the final manuscript.

## Institutional Review Board Statement

HINTS 7 data are publicly available, de-identified, and contain no individually identifiable information. This analysis of pre-existing, de-identified data is exempt from IRB review under 45 CFR 46.101(b)(4) [16]. No individual consent to participate was required.

## Informed Consent Statement

Not applicable. This study does not contain data from any individual person.

## Data Availability Statement

The datasets analysed during the current study are publicly available in the National Cancer Institute Health Information National Trends Survey (HINTS) repository [16, 29]. Analysis code is available from the corresponding author on reasonable request.

## Acknowledgements

Not applicable.

## List of Abbreviations

CI: Confidence interval (also Confident-Incorrect, contextdependent)
eHealth: Electronic health
GMM: Gaussian Mixture Model
HBV: Hepatitis B virus
HCV: Hepatitis C virus
HINTS: Health Information National Trends Survey
IRB: Institutional review board
NCI: National Cancer Institute
OR: Odds ratio
SD: Standard deviation

